# Extended Invariant Information Clustering is Effective for Leave-One-Site-Out Cross-Validation in Resting State Functional Connectivity Modelling

**DOI:** 10.1101/2021.06.07.21258389

**Authors:** Naoki Okamoto, Hiroyuki Akama

## Abstract

Herein, we propose a new deep neural network model based on invariant information clustering (IIC), proposed by Ji et al., to improve the modeling performance of the leave-one-site-out cross-validation (LOSO-CV) for a multi-source dataset. Our Extended IIC (EIIC) is a type of contrastive learning; however, unlike the original IIC, it is characterized by transfer learning with labeled data pairs, but without the need for a data augmentation technique. Each site in LOSO-CV is left out in turn from the remaining sites used for training and receives a value for modeling evaluation. We applied the EIIC to the resting state functional connectivity magnetic resonance imaging dataset of the Autism Brain Imaging Data Exchange. The challenging nature of brain analysis for autism spectrum disorder (ASD) can be attributed to the variability of subjects, particularly the rapid change in the neural system of children as the target ASD age group. However, EIIC demonstrated higher LOSO-CV classification accuracy for the majority of scanning locations than previously used methods. Particularly, with the adjustment of a mini-batch size, EIIC outperformed other classifiers with an accuracy >0.8 for the sites with highest mean age of the subjects. Considering its effectiveness, our proposed method might be promising for harmonization in other domains, owing to its simplicity and intrinsic flexibility. We are currently submitting this manuscript to Frontiers in Neuroinformatics.

## 1 Introduction

Machine learning discrimination has been widely applied to resting-state functional connectivity (RSFC) datasets which cover a wide range of neural diseases such as schizophrenia (Shen et al., 2010), mild cognitive impairment (Chen et al., 2016), and autism spectrum disorder (ASD) (Guo et al., 2017). Monk et al. (2009) demonstrated a difference in the strength of connectivity within the default mode network between ASD and control groups; the magnitude of this difference was correlated with the severity of symptoms. Guo et al. (2017) used a support vector machine (SVM) as a classifier and successfully differentiated between the two groups with >80% accuracy for data derived from a single imaging location in the Autism Brain Imaging Data Exchange (ABIDE) collection. The ABIDE is a dataset that consists of resting-state functional magnetic resonance imaging (MRI) images, structural images, and phenotypic data for 539 patients with ASD and 573 healthy developmental controls (TCs), recorded at 17 locations worldwide.

However, machine learning performs quite poorly on combined datasets derived from different imaging sites. This can be attributed to the effects of data heterogeneity, such as differences in the characteristics of MRI scanner suppliers, scan parameters (particularly the length of the repetition time [TR]), and the cohort setting, wherein variability in phenotype, specifically a deviation in the age and gender group composition, is unavoidable. Researchers commonly observe difficulty in harmonization with respect to a multi-source dataset mixture in the ABIDE analysis. Moreover, we experienced a considerable drop in the accuracy on excluding the overall data of each single site to create an independent testing set and featuring the remaining sites as the provider of the modeling set. This leave-one-site-out cross-validation (LOSO-CV) technique has certain complications, particularly in the case of ASD, such as the diverse age distribution for selecting the subjects. For example, Chen et al. (2016) used RSFC in two frequency bands and trained a discriminant model using feature selection and SVM. However, before running the LOSO-CV, they preselected the age bracket in the range of 12–18 years and removed the sites with <30 participants.

More recently, with an increasing interest in deep learning, researchers have proposed studies based on multi-layer network models for decoding by analyzing the static structure and dynamic modulation of RSFC. Heinsfeld et al. (2018) applied a denoising autoencoder model to the overall sites in the ABIDE and distinguished ASDs from TCs. The average accuracy was approximately 70% on evenly subtracting some portion of the data from all sites to leave the hybrid test set and train the model from the remaining subsets. However, they encountered a non-negligible fallen accuracy while distinguishing ASDs from TCs, upon changing the splitting methods for cross-validation. The average value remained 67% for the LOSO accuracy rates assigned to each of the left-out sites. Note that the ABIDE sites recording the largest accuracy decreases for LOSO compared to each of the single-site modeling results were those in which the mean ages were the highest or the lowest. The signs and symptoms of ASD differ across age groups. Moreover, growth and aging modulate the fundamental network architecture of RSFC (Supekar et al., 2009). The attention neural network (ANN) might be an alternative method to LOSO deep learning. Nonetheless, Niu et al. (2020) excluded five sites from the ABIDE dataset and supplemented the RSFC data with subject characteristics, such as gender and handedness.

In addition to the autoencoder (Hinton and Salakhutdinov, 2006), researchers also use adversarial generative networks for harmonizing MRI data (Yan et al., 2019) which adhere to the framework of contrastive learning (Ian et al., 2014). Furthermore, contrastive learning together with the data augmentation technique have been adopted to overcome the individual variability and noise in MRI experiments which cause a failure in machine learning. Contrastive learning is a self-supervised method that takes advantage of a conjugated data pair sampled from the mixing set of different categories. The pair of similar category generates similar output and that of the different one generates dissimilar output to grasp the category essence. For example, Ji et al. (2019) proposed invariant information clustering (IIC) as a discriminant model that learns noise-independent features from pairs of original images and noise-added ones. The IIC has been devised to minimize mutual information loss for paired data samples. The IIC, despite its simplicity, usability, and potentiality remains, however, in the early stage of applicability to domains other than simple image discrimination, which was its original target. Beyond this target realm, the input modeling level needs to be modified to introduce the IIC into the ABIDE fMRI dataset, as it is characterized by less contrastive features and low frequency signal oscillation.

In the wake of previous studies pertaining to the LOSO-oriented ABIDE analysis, we proposed an IIC-based deep neural network model based on the original IIC of Ji et al. This model was developed for discriminating the labeled data of ASD and TC in the ABIDE dataset only with the RSFC data obtained from almost all imaging locations. Our bias regulation technique, the extended-IIC (EIIC), consisted of transfer learning with labeled data concatenated with contrastive prior learning where the framework was borrowed from the original IIC. Despite its efficiency, the original IIC requires data augmentation, such as image rotation and flipping, so that it cannot be simply applied to datasets where this technique affects signal quality, since human intuition-based manual feature extraction is by nature difficult to perform, as with RSFC analysis.

However, we improved the IIC by the following steps: a data pair preparatory step with labelled information that precisely substitutes the data augmentation technique and is useful for the contrastive learning of this type of datasets, if posterior supervised learning is provided subsequently to the original IIC to take over the learned weights conveying the intrinsic discriminant information. From a modeling perspective, we examined the decoding feasibility and predictive power of the EIIC by executing LOSO-CV for the sites of the ABIDE dataset. In addition, we used several algorithms to evaluate the relative fit of the models in terms of the effectiveness in harmonizing multi-data sources of the ABIDE through LOSO. The classifiers were learned following the extraction of the blood oxygenation level dependent (BOLD) time-course series from each region of interest (ROI) of the Harvard-Oxford (HO) brain atlas (Desikan et al. 2006). We computed traditional classifiers, such as the support vector machine (SVM) and random forest (RF) as baselines, which were compared with the performances of the EIIC. We were also able to identify the best method to adjust for the issues of LOSO, specifically weakness to the age distribution bias, to obtain a desirable precision superior to previously published results.

## 2 Methods

This study was performed in accordance with the tenets of the Declaration of Helsinki and was approved by the Ethics Committee of the Tokyo Institute of Technology (approval number: 2019091).

### 2.1 Datasets

The ABIDE is available as preprocessed data using a pipeline software. “ABIDE Preprocessed” (Craddock et al. 2013) releases numerous features for analysis, including regional homogeneity, the amplitude of low-frequency fluctuation, and average time series data for ROIs of seven different brain atlases. In this study, we used the average time series of the HO brain atlas, preprocessed with a configurable pipeline for the analysis of connectomes (C-PAC) (Craddock et al. 2013).

### 2.2 An outline of the proposed method

The original IIC is a neural network model developed to perform image discrimination and segmentation tasks without labels. IIC models are trained to maximize the mutual information between outputs that assign classes to paired data. An IIC is able to learn a model that assigns one data item to each class with high probability, while avoiding meaningless assignments, such as attributing all data to a single class. In addition, the IIC has an overclustering (oc) head, which refers to an output that assigns more classes than expected, such that it can handle data comprising severe noise and unexpected classes. According to the IIC of Ji et al. (2019), the paired data for computing mutual information comprises an original image and its randomly perturbed version.

Data augmentation refers to the process of creating a transformed image from unlabeled data, in the context of self-supervised models by contrastive learning. It generally involves transforming the actual data to create one that resembles the original data. However, we assumed that the actual data in the ABIDE collection was already augmented for contrastive learning by the addition of several noises to the unknown functional connectivity patterns, typical of ASD and TC. Based on this interpretation, the data pairs were produced according to the aforementioned labels for prior self-supervised learning.

EIIC is characterized by an assembly line network that enables the double-purpose mechanism (paired or independent processing), which in turn can be summarized as a two-step process for transfer learning. The first step involves prior learning, which can ignore the output for discrimination. However, it trains the entire network using only the IIC loss (described below) of the two outputs as follows: one for the pair with similar or different labels and the other for overclustering. Following a sufficient number of epochs for prior learning, we fixed the values of all weights except those in the layer immediately before the output, such that no further training was performed at this stage. Posterior training is the core part of transfer learning, which trains only the penultimate layer using the CE loss (described below) for discrimination. Therefore, the sub-model up to this layer will likely be sufficiently robust to noise using the IIC framework. Moreover, we will train the final model for discrimination based on the established learning results.

### 2.3 Details of the proposed method

Figure 1 provides an overview of the above-mentioned EIIC **(A:** prior learning; **B**: transfer learning). The overall flow of analysis, common to all classifiers, was as follows. First, we applied the Fisher’s z-transformation to the functional connectivity matrix (size, 110 × 110), derived from the standardized ROI-averaged BOLD time series data in “ABIDE Preprocessed.” We also extracted the lower triangular part of the matrix as the input vector of 5,995 elements for the subsequent analysis. We excluded any “NaN” for the BOLD in some ROIs of the HO atlas, thereby omitting the Carnegie Mellon University (CMU) as a critical site. This is because only one subject was left following the screening process.

**Figure 1.**
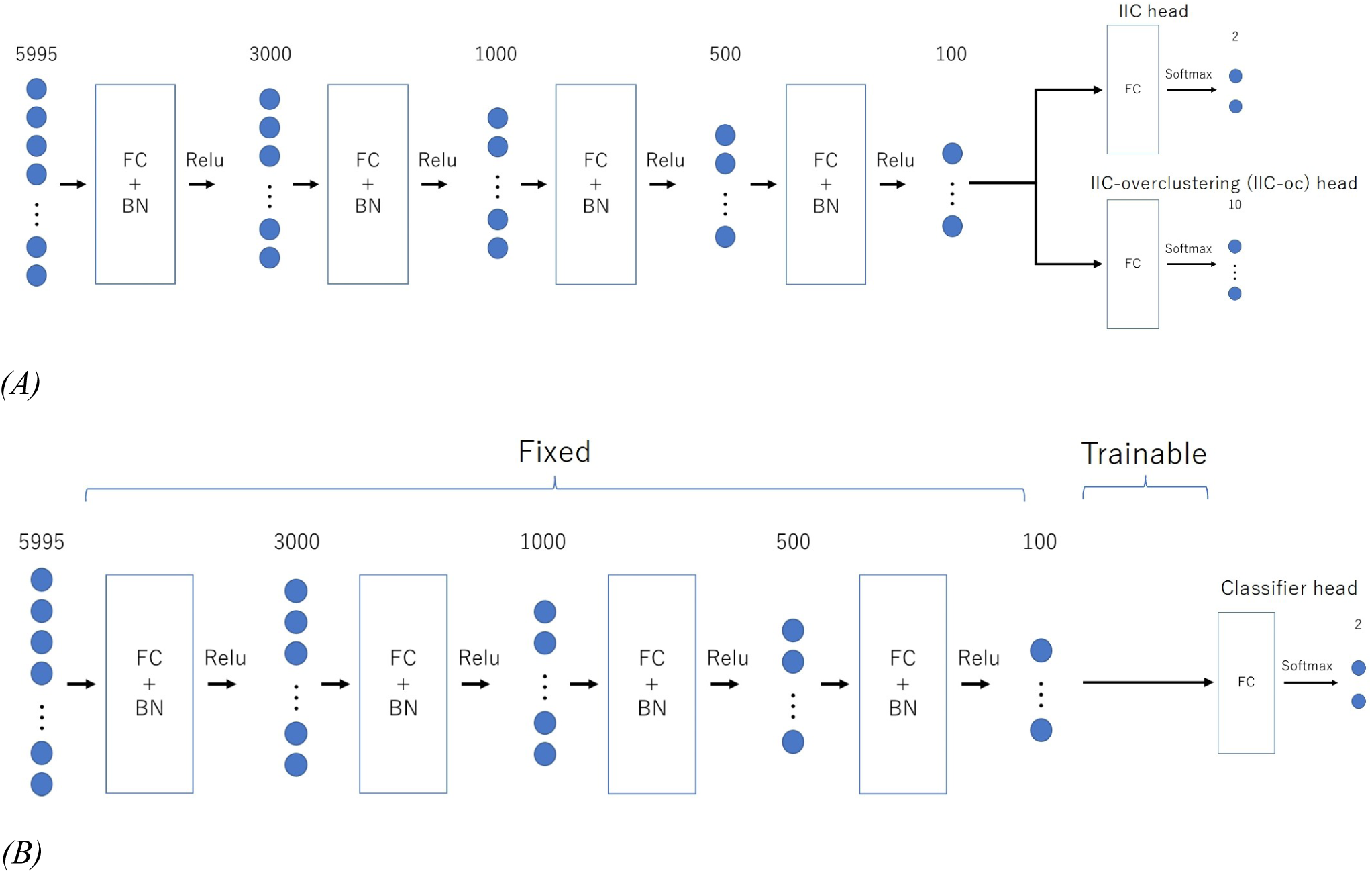
Architecture of the two-step EIIC mechanism. (A): contrastive prior learning (simple IIC structure) for the first step; (B): transfer learning with IIC for the second step. IIC: invariant information clustering. EIIC: extended invariant information clustering.

The basic EIIC architecture is a combination of a fully connected (FC) layer and a batch normalization (BN) layer (Ioffe and Szegedy, 2015). While the FC layer adopts a linear sum over all nodes in the previous one, the BN layer standardizes the input for each mini-batch in mini-batch learning. The BN layer is widely used to facilitate learning by preventing internal covariate shifts among the outputs of each layer. For the activation function, the softmax function is only used for the output layer, in contrast to the ReLu function used for the remaining four intermediate layers.

We used a combination of the following two types of loss functions for training the model: (i) IIC loss, which aimed to maximize the mutual information content of the output between paired data; and (ii) cross-entropy loss, designed to generate larger output corresponding to the class of correct labels for each data. In IIC learning, the mutual information content of the output Φ(x) from the paired data (x, x′) is maximized and expressed in the following formula:

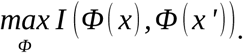

The output of the neural network can be regarded as the distribution of the probability of being assigned to each class if the activation function of the output layer is a softmax function. By performing a marginalization of the training batch, the simultaneous probability P of the class assignment for a pair of data can be expressed as follows:

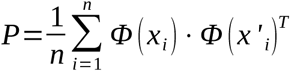

The equation assumes the form of a matrix of C×C, where C is the number of classes. The sum of the rows or columns of this matrix yields the following:

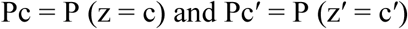

Moreover, the mutual information can be formulized as follows:

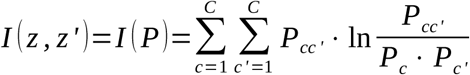

Ji et al. (2019) proposed the efficacy of a coefficient λ that adds weight to the marginal entropy maximization term for the aforementioned mutual information content. For the training, we set the value of λ to 5.0 for EIIC in the following formula, where H(z) represents the marginal entropy of the variable z:

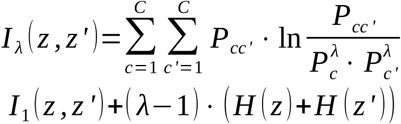

While training the model, we used the mutual information introduced by λ multiplied by -1 as the IIC loss, following a division by the natural logarithm of the number of discriminant classes. This helped us to transform the problem of maximizing the amount of information to that of minimizing the loss function. Furthermore, in the case of a two-class discrimination, 1 - Φ(x) can be assumed as the probability distribution for the class opposite to the one that comprises the predicted x. We used the cross entropy loss (CE-loss), i.e., 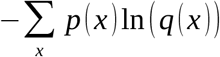, as the loss function for discriminating the two classes. We first trained the IIC head and IIC-oc head by summing the IIC losses (pre-training). Moreover, we trained the classifier head by CE-loss, fixing all weights except those directly connected to the output layer (transfer learning) (Figure 2). In summary, the loss functions of the EIIC for each training phase were as follows:

· Prior learning

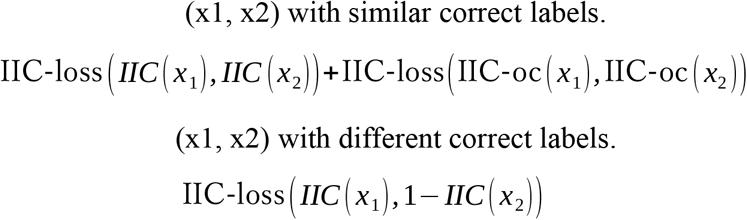

· Transfer learning

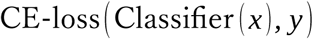

For the above-mentioned models, the optimization method comprised mini-batch learning with Adam (Kingma et al., 2014). Mini-batch learning uses a portion of the training data at one time, instead of inputting all training data into the model at once. We defined the update of weights by a mini-batch as one “iteration.” Moreover, we attempted using different batch sizes of 500, 3,000, and 5,000 at one time. The update of weights was executed in accordance with the sequential random extraction of mini-batches from the training data. We regarded one “epoch” as terminated upon our failure to extract the mini-batch. Please refer to the Supplementary Material for details on the process of updating the weights.

**Figure 2.**
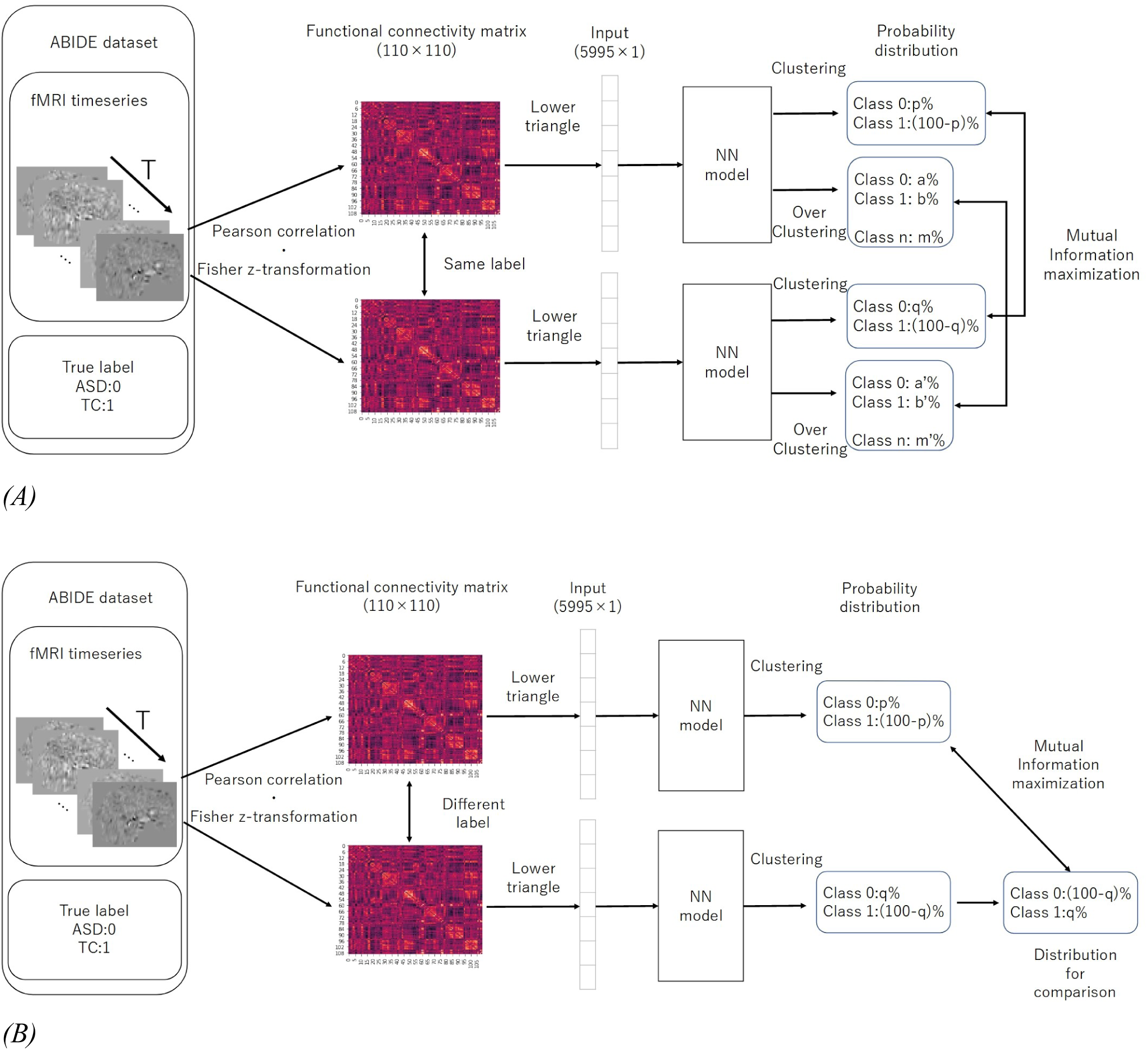
Computation of IIC-loss for pairs with similar (A) and different (B) correct labels. IIC: invariant information clustering. The multi-class is assumed at the overclustering head output. In our method, data pairs for contrastive learning were based on single data labels instead of with a data augmentation technique.

### 2.4 Evaluating the Accuracy (LOSO-CV)

We executed LOSO-CV as a splitting strategy, for which the data with a similar imaging location was considered a subpopulation. This facilitated testing the performance of the models. The data of each site was omitted as a test set to be used for computing the classification accuracy (Figure 3). The remaining sites were split into the training and validation sets and employed for learning with an additional fivefold CV to ensure an early stopping of the training. Considering the balance of ASD and TC subjects, the composition of the imaging locations in each of the five folds would be similar to the entire training data to the maximum possible extent. Herein, we used one split-out fold as the validation set and the rest as the training set, by changing the roles in turn.

**Figure 3.**
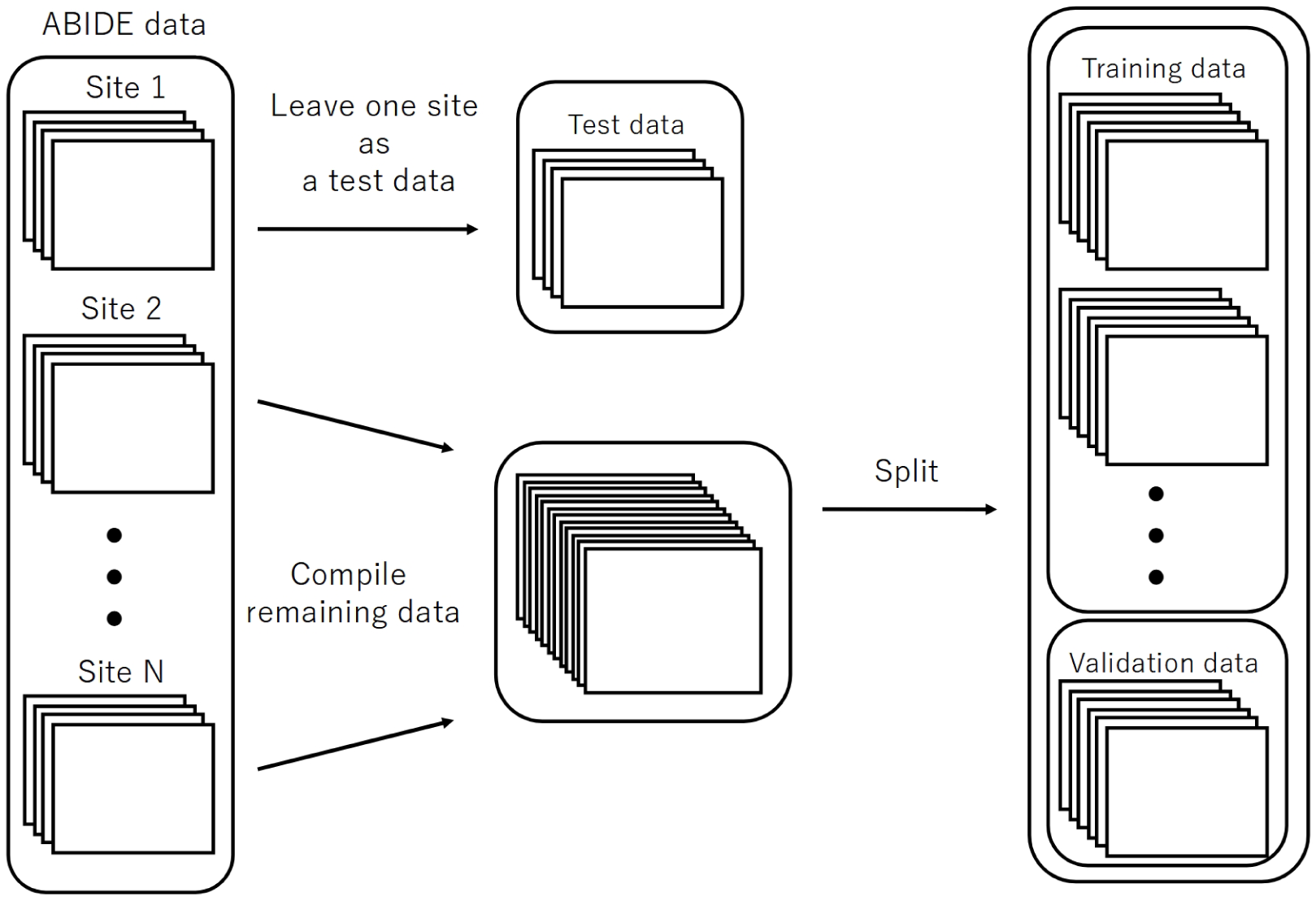
The splitting method for the evaluation of LOSO-CV accuracy. LOSO-CV, leave-one-site-out cross-validation.

For EIIC, we updated the parameters of the model using the training set until the loss function for a validation set did not further decrease within the predetermined number of epochs for early stopping of the training. The model with the smallest loss function for the validation set was determined to be trained. As the loss function for the validation, we adopted the sum of the IIC head and IIC-oc head losses for the paired input data for the pre-training. Five trained models were generated, corresponding to five separate validation sets. Inspired by the basic idea of ensemble learning, we inputted the test data to each of the five validated models to obtain an average of the two output nodes (corresponding to the category of 0 or 1, respectively) at the classifier head of transfer learning. We subsequently adopted the mean value of these five outputs to decide if the predicted category was 0 or 1 for the corresponding input by rounding off the value. The input for each model was standardized using the mean and standard deviation of the training data, corresponding to the model.

In relation to SVM and RF, we ran the dimensionality reduction by the principal component analysis (PCA) as well as the adjustment of hyperparameters through the grid search nested into CV. The codes for EIIC employed Pytorch (Paszke et al., 2019). In contrast, those for SVM and RF were implemented using the Python package scikit-learn (Pedregosa et al., 2011) (all available at: https://github.com/nokamot/EIIC).

## 3 Results

Table 1 summarizes the discrimination accuracy of LOSO for the three classifiers, i.e., EIIC, SVM, and RF. The accuracy rates of the EIIC were obtained with the smallest mini-batch size of 500. The aforementioned parameter setting provided the best score compared with the larger sizes (for those of 3,000 and 5,000, refer to the tables in the Supplementary Materials). EEC elicited significantly higher accuracy than other classifiers. Moreover, the EIIC recorded >80% accuracy and outperformed other classifiers for the sites with highest mean age of the subjects (CALTECH: 0.81 for 26.5, SBL: 0.84 for 35.2, and USM: 0.80 for 23.7) except for MAX_MUN (0.55 for 27.9). The classifier did not improve the discrimination accuracy for the low age group, which in turn remained flat regardless of any hyperparameter setting (KKI, OHSU, and STANFORD). However, the EIIC was successful in providing better models to the sites shifted to infancy (SDSU: 0.79 vs. 0.63 for 14.2, UCLA: 0.72 vs. 0.66 for 13.3, UM: 0.70 vs. 0.64 for 14.5, and YALE: 0.70 vs. 0.65 for 12.8) than the auto-encoder model of Heinsfeld et al.

**Table 1.**
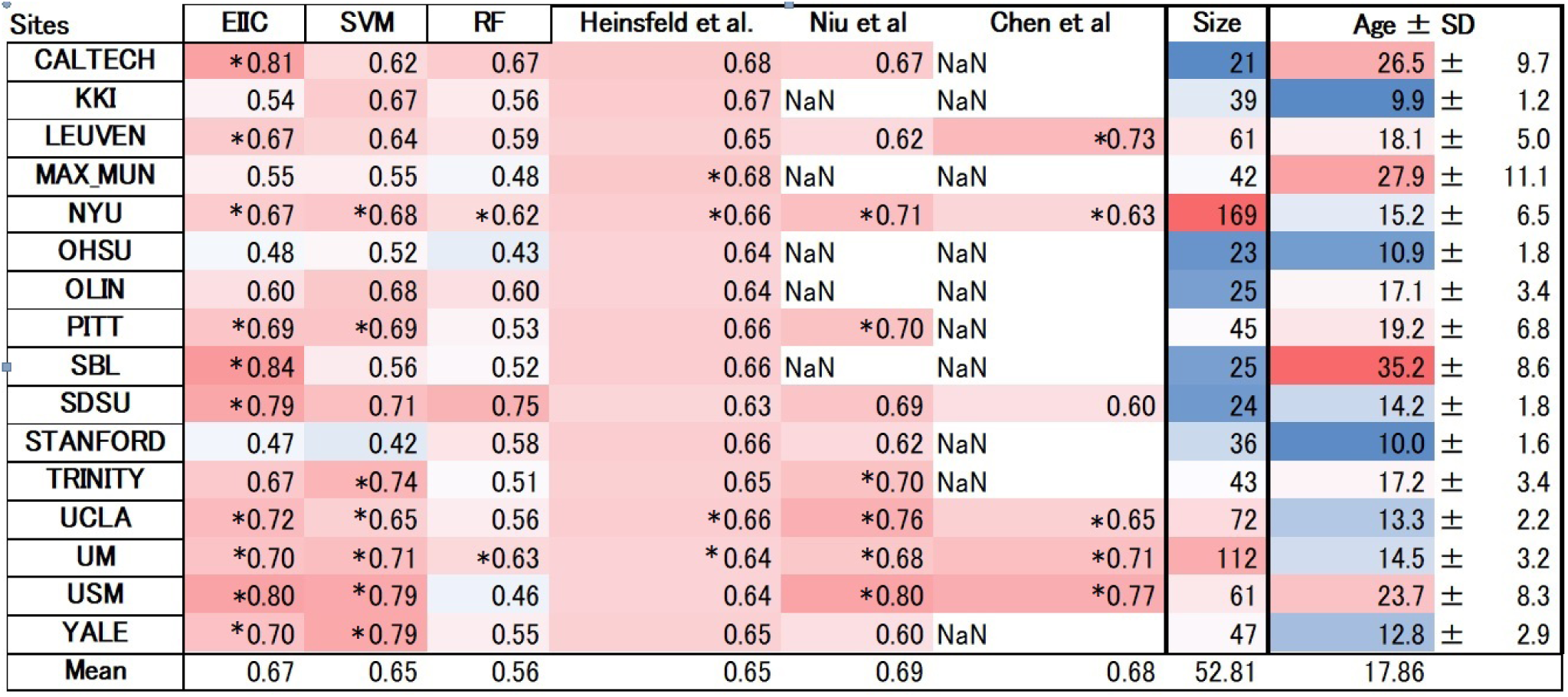
The LOSO-CV accuracy for each classifier and characteristics of each ABIDE site. We adjusted the background color based on cell value. The result of each classifier (column) is depicted darker in red if an accuracy rate > 0.5 approaches 1 (the perfect value). In contrast, values <0.5 acquired a darker background shade of blue by approaching 0 (the complete misclassification). The dataset size and the mean participants’ age are also emphasized by a similar gradual color scale between darkest red (maximum value) and darkest blue (minimum value). *: p < 0.01, binomial test, chance level set at 50%. LOSO-CV, leave-one-site-out cross-validation.

## 4 Discussion

ASD is a developmental disorder that occurs in early childhood and undergoes temporal variation in RSFC. This necessitates a uniform age group for the subjects. However, the condition makes it difficult to collect large-scale data. Therefore, some member organizations of ABIDE were biased towards a particular age group, which complicated and led to the occasional failure of LOSO-CV. EIIC succeeded in recording accuracy values >80% for sites that primarily comprised older subjects (>20 years), namely the CALTECH, SBL, and USM, except for MAX-MUN. The low discriminative power revealed in MAX-MUN might be attributable to the low temporal resolution with the length of repetition time (TR) set to 3,000 ms. The aforementioned sites specific to the relatively elderly generation elicited the largest accuracy drop for LOSO, compared to each within single-site modeling in the denoising autoencoder of Heinsfeld et al. The sample sizes of CALTECH and SBL were quite small (21, 25). Thus, the EIIC was essentially robust because of the scarcity of data. However, the number of subjects was larger than 60 in the USM that recorded 80% accuracy with a mean age of 23. Hence, it would be somewhat unreasonable to impute the effectiveness of EIIC only based on its robustness to the condition of data paucity.

Compared to the above-mentioned sites, the results were more conflicting with respect to studies that recruited younger children. Of the sites with a mean age <18 years, six sites provided higher accuracy to EIIC than the methods proposed by Heinsfeld et al. However, the latter outperformed our methods for the following sites: KKI, 0.54 vs. 0.67 for 9.9; OHSU, 0.48 vs. 0.64 for 10.9; and STANFORD, 0.47 vs. 0.66 for 10.0 (displayed in order of the site name, the accuracy of EIIC vs. that of the Denoising Autoencoder, and the mean site age). The graphical index of RSFC is significantly different between children aged 7–9 years and those aged 19–22 years (Supekar et al., 2009). Furthermore, small children generate more noise because of body motion in the MRI system (Hull et al., 2016), which may exert a significant effect on the data quality. Hence, composite factors should be considered for the three sites, in terms of the generalization capability of the EIIC. OHSU adopted a relatively long TR set to 2,500 ms. In addition, it provided extremely small volumes (82 vols) for each functional scan. Therefore, similar argument for MAX-MUN might apply to this site. Moreover, an inter-vendor harmonization might be required since STANFORD employs a GE MRI scanner, which belongs to a minority site group.

An unexpected finding is that SVM achieved accuracy comparable with the deep learning models, despite being known as a classical machine learning strategy. This might be attributed to the dimensionality reduction by PCA and the hyperparameter adjustment by grid-search, notwithstanding its lower effectiveness for RF. However, we can underscore the advantage of EIIC over SVM, particularly for sites with higher age groups. We identified this advantage on setting the size of the mini-batch to 500. The effectiveness of the small mini-batch size was possibly because the learning number of times within one epoch increased as it was defined as the proportion of data size, with respect to the mini-batch size. In other words, the larger mini-batch size that reduced the learning number of times might result in an inability to escape from the local solution in which the learning process is trapped (see the Supplementary Material). However, further optimization of the mini-batch size is required to fuel future research.

In sum, the novelty of our methods lies in the fact that we concatenated two independent processes, i.e., contrastive prior learning and transfer learning. Therefore, we were able to propose a solution for the most challenging issues in brain MRI modelling; specifically, RSFC patterns which are hard to distinguish between ASD and TC subjects if they visited different sites for their scans. However, despite the considerable merit of our modeling strategy, EIIC has some limitations, some of which may be worthy of future inquiry. As noted above, EIIC might be less suited to predicting the attribution for cohorts of younger children. Furthermore, it remains to be specified how we will improve the loss function for the pairs of different labels in contrastive learning. It may be that these issues are connected with each other, which will lead us to unveil the most efficacious method for multi-site modeling in ASD research.

## 5 Conclusion

In conclusion, we proposed a new algorithm for multi-site harmonization of an RSFC dataset, derived from different sources. EIIC, as an extension of the original IIC, is a type of contrastive learning. EIIC achieved good generalization performance in the classification of unknown sites by the new application of LOSO-CV to the ABIDE data collection. The classification accuracy following EIIC application was higher for the majority of locations than previously used methods. The deep neural network used in this study was relatively simple, with only one sequence of an FC layer and a batch regularization layer. However, there is sufficient room for improvement based on data augmentation or other machine learning strategies by using the sliding window approach for the dynamic functional connectivity (DFC), or extending the maximization of mutual information to multiple categories to be discerned. Considering the present effectiveness, our proposed method might be promising for harmonization in other domains, owing to its simplicity and intrinsic flexibility. We believe that it will be also effective for multi-source─but relatively small-sized─datasets, and will achieve optimal efficacy when the data augmentation technique does not affect the signal quality.

## 7 Author Contributions

Conceptualization: NO and HA; data curation and analysis: NO and HA; investigation: NO and HA; methodology: NO and HA; supervision: HA; and writing: NO and HA. All authors contributed to manuscript revision, read, and approved the submitted version.

## 9 Abbreviations

RSFC: resting-state functional connectivity;
ASD: autism spectrum disorder;
DMN: default mode network;
ABIDE: Autism Brain Imaging Data Exchange;
LOSO: leave-one-site-out;
SVM: support vector machine;
IIC: invariant information clustering;
EIIC: extended invariant information clustering;
HO: Harvard-Oxford;
BOLD: blood-oxygenation-level-dependent;
FC: full coupling;
BN: batch regularization;
CV: cross-validation;
RF: random forest.

## 10 Acknowledgments

We thank Editage (www.editage.com) for their English language editing service.

## 11 Data Availability Statement

The datasets generated for this study can be accessed by the public. (http://preprocessed-connectomes-project.org/abide/index.html)

## Supplementary Material

### 1 Supplementary Data

#### I. A comparison between the EIIC and another contrastive learning model

Our proposed EIIC method handles label pairs in the same way as the method proposed by Ktena et al. (2018). The latter is an example of a contrastive learning model, applied to the ABIDE dataset. Ktena et al. used a graph convolutional neural network, which reflected the information regarding brain structure in the RSFC data to train their model with a distance index. This helped to discriminate between AD and TC cases. In contrast, EIIC has the advantage that the output of paired learning is not a distance within a pair, but rather a model for clustering itself. Therefore, new incoming data can be directly labeled without making a pair with an existing one.

#### II. Mutual Information

The mutual information I of two random variables X and Y can be calculated using Equation (1) to evaluate the degree of dependence between the two random variables:

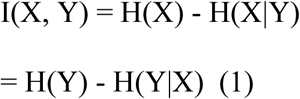

The mutual information refers to the difference between these two terms, which implies the extent to which knowing one of the two random variables can reduce the uncertainty about the other. The desired maximization of I(x,y) is achieved by minimizing and maximizing the conditional entropy H(X|Y) and marginal entropy H(X), respectively. The conditional entropy is minimized when X and Y are completely predictable from one to the other. Thus, it is optimal to assign data to one class with high probability. Considering that the marginal entropy is the uncertainty of X or Y itself, maximization avoids undesirable solutions that would assign data to a single class regardless of the label.

#### III. Weight updates

The definition of one “epoch” was somewhat unusual while training the EIIC model because of the alternation as an epoch unit that we first updated the weights for similar label pairs thoroughly, and then for different labels. The optimization method determines the procedure to update the weights based on the gradient of the loss function, calculated by an error back propagation. Moreover, Adam is a widely used method in neural network training.

The formula for the the update of the weight w is given below:

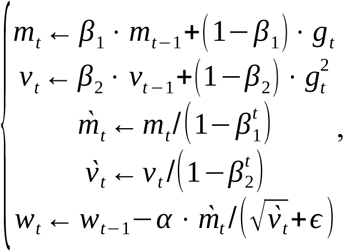

w_t_ denotes the weight updated t times and g_t_ is the gradient of the loss function for w_t-1_. The learning coefficient α and those for the moving average of the gradients β1 and β2 are hyperparameters to be determined in advance. We set β1=0.9 and β2=0.999 for both models. We assigned the learning coefficient α, such that it oscillated every epoch as α_t_ according to the SGDR algorithm (Loshchilov and Hutterm, 2016), with a maximum value of α_max_=0.0001.

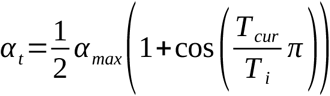

Initially, Tcur was set to 0 and T_i_ was set as a positive integer as a hyperparameter. With an increase in T_cur_ by 1 with each learning step, the learning coefficient α_t_ approaches 0. Upon reaching 0, T_i_ is updated and T_cur_ gets initialized to 0. The initial value of T_i_ was set to 2 and increased by 2 on each update. Through this operation, we could expect the learning coefficient to increase rapidly upon updating T_i_. It assists the model in escaping from a local solution.

#### IV. Adjusting the mini-batch size

The tables highlight the results obtained with mini-batch sizes of 3,000 and 5,000.

#### V. Data and scripts

The data and the scripts are all available at: https://github.com/nokamot/EIIC

This is a repository of codes for An Extended Invariant Information Clustering, effective for the LOSO-CV in resting-state functional connectivity modeling.

a) Requirements

We used a docker container of pytorch/pytorch:1.8.1-cuda11.1-cudnn8-runtime for analysis. In addition, scikit-learn, pandas, and openpyxl are required.

b) How to run

### 1. Download data

Download the preprocessed ROI time series of the ABIDE dataset and phenotypic data from the download page of ABIDE Preprocessed.

### 2. Set config file

Prepare two directories to save the intermediate files (input data and labels) and final output (trained models).

Edit four relative path items of param_set.py as follows:

i. source_dir: Directory of intermediate files
ii. output_dir: Directory of result files
iii. preparation_params[‘label_file_path’]: Phenotypic data
iv. preparation_params[‘path_structure’]: Each ROI timeseries files

### 3. Train models and output results

Pull the Pytorch docker image.

#### sudo docker pull pytorch/pytorch:1.8.1-cuda11.1-cudnn8-runtime

Start the docker container mounting a directory, including these codes and necessary files, as mentioned in Section 2 (Set config file).

**sudo docker run -it --rm --gpus device=0 -v /Path/to/codes/in/host:/Path/to/codes/in/container pytorch/pytorch:1.8.1-cuda11.1-cudnn8-runtime**

Install the required packages by pip and run in the container.

**pip install scikit-learn pandas openpyxl**

**cd /Path/to/codes/in/container**

**python run.py**

**2 Supplementary Table.**

| SITES | EIIC | SVM | RF | Heinsfeld et al. | Size | Age ± SD |
| --- | --- | --- | --- | --- | --- | --- |
| CALTECH | 0.76 | 0.62 | 0.67 | 0.68 | 21 | 26.5 ± 9.7 |
| KKI | 0.44 | 0.67 | 0.56 | 0.67 | 39 | 9.9 ± 1.2 |
| LEUVEN | 0.62 | 0.64 | 0.59 | 0.65 | 61 | 18.1 ± 5.0 |
| MAX MUN | 0.48 | 0.55 | 0.48 | 0.68 | 42 | 27.9 ± 11.1 |
| NYU | 0.63 | 0.68 | 0.62 | 0.66 | 169 | 15.2 ± 6.5 |
| OHSU | 0.52 | 0.52 | 0.43 | 0.64 | 23 | 10.9 ± 1.8 |
| OLIN | 0.72 | 0.68 | 0.60 | 0.64 | 25 | 17.1 ± 3.4 |
| PITT | 0.64 | 0.69 | 0.53 | 0.66 | 45 | 19.2 ± 6.8 |
| SBL | 0.76 | 0.56 | 0.52 | 0.66 | 25 | 35.2 ± 8.6 |
| SDSU | 0.67 | 0.71 | 0.75 | 0.63 | 24 | 14.2 ± 1.8 |
| STANFORD | 0.58 | 0.42 | 0.58 | 0.66 | 36 | 10.0 ± 1.6 |
| TRINITY | 0.63 | 0.74 | 0.51 | 0.65 | 43 | 17.2 ± 3.4 |
| UCLA | 0.74 | 0.65 | 0.56 | 0.66 | 72 | 13.3 ± 2.2 |
| UM | 0.71 | 0.71 | 0.63 | 0.64 | 112 | 14.5 ± 3.2 |
| USM | 0.75 | 0.79 | 0.46 | 0.64 | 61 | 23.7 ± 8.3 |
| YALE | 0.62 | 0.79 | 0.55 | 0.65 | 47 | 12.8 ± 2.9 |
| Mean | 0.64 | 0.65 | 0.56 | 0.65 |  |  |

Mini-batch size: 5,000
| SITES | EIIC | SVM | RF | Heinsfeld et al. | Size | Age ± SD |
| --- | --- | --- | --- | --- | --- | --- |
| **CALTECH** | 0.62 | 0.62 | 0.67 | 0.68 | 21 | 26.5 ± 9.7 |
| **KKI** | 0.69 | 0.67 | 0.56 | 0.67 | 39 | 9.9 ± 1.2 |
| **LEUVEN** | 0.59 | 0.64 | 0.59 | 0.65 | 61 | 18.1 ± 5.0 |
| **MAX MUN** | 0.60 | 0.55 | 0.48 | 0.68 | 42 | 27.9 ± 11.1 |
| **NYU** | 0.64 | 0.68 | 0.62 | 0.66 | 169 | 15.2 ± 6.5 |
| **OHSU** | 0.43 | 0.52 | 0.43 | 0.64 | 23 | 10.9 ± 1.8 |
| **OLIN** | 0.72 | 0.68 | 0.60 | 0.64 | 25 | 17.1 ± 3.4 |
| **PITT** | 0.51 | 0.69 | 0.53 | 0.66 | 45 | 19.2 ± 6.8 |
| **SBL** | 0.56 | 0.56 | 0.52 | 0.66 | 25 | 35.2 ± 8.6 |
| **SDSU** | 0.67 | 0.71 | 0.75 | 0.63 | 24 | 14.2 ± 1.8 |
| **STANFORD** | 0.56 | 0.42 | 0.58 | 0.66 | 36 | 10.0 ± 1.6 |
| **TRINITY** | 0.58 | 0.74 | 0.51 | 0.65 | 43 | 17.2 ± 3.4 |
| **UCLA** | 0.58 | 0.65 | 0.56 | 0.66 | 72 | 13.3 ± 2.2 |
| **UM** | 0.67 | 0.71 | 0.63 | 0.64 | 112 | 14.5 ± 3.2 |
| **USM** | 0.75 | 0.79 | 0.46 | 0.64 | 61 | 23.7 ± 8.3 |
| **YALE** | 0.60 | 0.79 | 0.55 | 0.65 | 47 | 12.8 ± 2.9 |
| **Mean** | 0.61 | 0.65 | 0.56 | 0.65 |  |  |

**3 Supplementary Figures.**
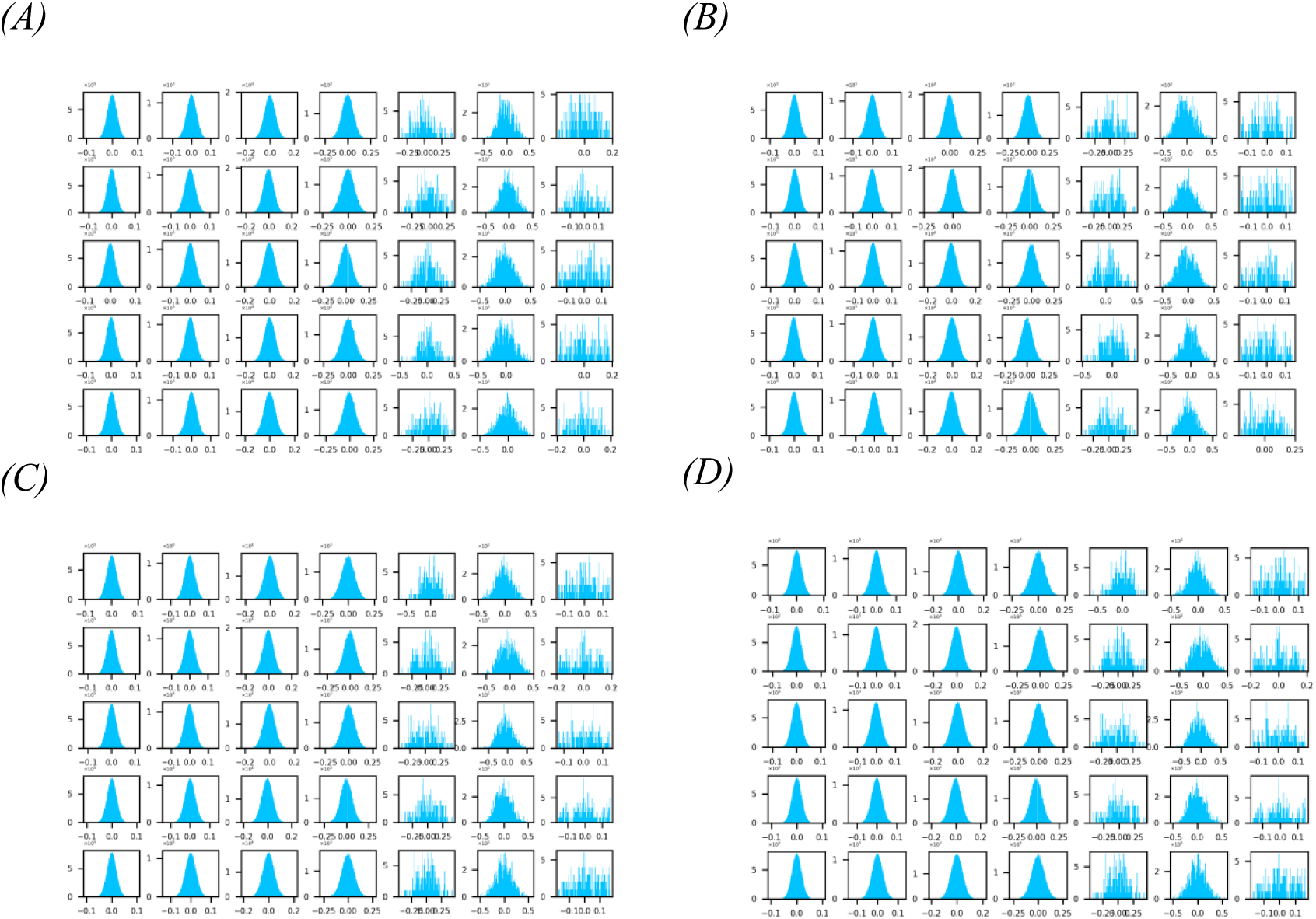

Four typical examples of the site-wise histograms of the fully connected layers’ weights in the EIIC computation with the number of bins set to 100. These are A:SBL(0.84), B:CALTECH(0.81), C:OHSU(0.48), and D:STANFORD(0.47), which are four organizations providing source data recording the two highest and two lowest accuracy rates. All the histograms for the 16 organizations in total (“hist_300dpi.zip”) and the Python code for computation (“check_weights.ipynb”) are published and available at: https://github.com/nokamot/EIIC. The numerical data for the overall layers’ weights will be provided upon the request of readers under the form of “result_models_bs500_<site>.bf”, of which the file size is approximately 800 Mb. In each histogram, the five rows correspond to each fold of the 5-fold cross-validation, whereas the seven columns correspond to each layer in the way that they lined up in the progress order from the left to the right. The leftmost four columns in each histogram represent the layers in the contrastive prior learning (simple IIC structure) for the first step (Figure 1), of which the weights are fixed and inherited to the transfer learning for the second step. These histograms for the fully connected layers took a sort of bell shape similar to the normal distribution around 0, regardless of the resulted classification accuracy. Conversely, the rightmost three columns in each histogram stand for the output layers, including IIC head, IIC_oc head (Figure1 A for the prior learning), and Classifier_head (Figure1 B for the posterior learning), respectively. When comparing the weight distributions of the IIC head (third from the right) and the Classifier head (the far right) sharing the same number of output nodes, it turned out that for the majority of the data source sites, the standard deviation of the weights at the latter was reduced to almost half of that at the former, signifying that the final discrimination in the posterior learning was based on the whole output from the proximate layer without reflecting the influence of some particular nodes there.

## References

Chen, H., Duan, X., Liu, F., Lu, F., Ma, X., Zhang, Y., Uddin, L. Q., Chen, H. (2016). Multivariate classification of autism spectrum disorder using frequency-specific resting-state functional connectivity—A multi-center study. Prog. Neuropsychopharmacol. Biol. Psychiatry 64, 1–9. doi: 10.1016/j.pnpbp.2015.06.014

Chen, X., Zhang, H., Gao, Y., Wee, C.-Y., Li, G., Shen, D. (2016), High-order resting-state functional connectivity network for MCI classification. Hum. Brain Mapp. 37, 3282–3296. doi: 10.1002/hbm.23240

Craddock, C., Benhajali, Y., Chu, C., Chouinard, F., Evans, A., Jakab, A., Khundrakpam, BS., Lewis, JD., Li, Q., Milham, M., Yan, C., Bellec, P. (2013). The Neuro Bureau Preprocessing Initiative: open sharing of preprocessed neuroimaging data and derivatives. Front. Neuroinform. Conference Abstract: Neuroinformatics 2013. doi: 10.3389/conf.fninf.2013.09.00041

Craddock, C., Sikka, S., Cheung, B., Khanuja, R., Ghosh, SS., Yan, C., Li, Q., Lurie, D., Vogelstein, J., Burns, R., Colcombe, S., Mennes, M., Kelly, C., Di Martino, A., Castellanos, FX., Milham, M. (2013). Towards Automated Analysis of Connectomes: The Configurable Pipeline for the Analysis of Connectomes (C-PAC). Front. Neuroinform. Conference Abstract: Neuroinformatics 2013. doi:10.3389/conf.fninf.2013.09.00042

Desikan, RS., Ségonne, F., Fischl, B., Quinn, BT., Dickerson, BC., Blacker, D., Buckner, RL., Dale, AM., Maguire, RP., Hyman, BT., Albert, MS., Killiany, RJ. (2006) An automated labeling system for subdividing the human cerebral cortex on MRI scans into gyral based regions of interest. Neuroimage 31(3):968–980

Guo, X., Dominick, K.C., Minai, A.A., Li, H., Erickson, C.A., Lu, L.J. (2017). Diagnosing autism spectrum disorder from brain resting-state functional connectivity patterns using a deep neural network with a novel feature selection method. Front. Neurosci. 11, 460. doi: 10.3389/fnins.2017.00460

Heinsfeld, A.S., Franco, A.R., Craddock, R.C., Buchweitz, A., Meneguzzi, F. (2018). Identification of autism spectrum disorder using deep learning and the ABIDE dataset. Neuroimage Clin. 17, 16– 23. doi: 10.1016/j.nicl.2017.08.017

Hinton, G.E., and Salakhutdinov, R.R. (2006). Reducing the dimensionality of data with neural networks. Science 313, 504–507. doi: 10.1126/science.1127647

Hull, J.V., Dokovna, L.B., Jacokes, Z.J., Torgerson, C.M., Irimia, A., Van Horn, J.D. (2017). Resting-state functional connectivity in autism spectrum disorders: A review. Front. Psychiatry 7, 205. doi: 10.3389/fpsyt.2016.00205

Ian, J.G., Jean, P.A., Mehdi, M., Bing, X., David W.F., Sherjil, O., Aaron, C., Yoshua, B. (2014). Generative adversarial networks. 1406.2661. https://arxiv.org/abs/1406.2661

Ioffe, S., and Szegedy, C. (2015). Batch normalization: Accelerating deep network training by reducing internal covariate shift. In Francis Bach and David Blei, editors, Proceedings of the 32nd International Conference on Machine Learning, vol. 37 of Proceedings of Machine Learning Research, pp. 448–456, Lille, France.

Ji, X., Henriques, J.F., Vedaldi, A. (2019). Invariant information clustering for unsupervised image classification and segmentation. In Proceedings of the IEEE/CVF International Conference on Computer Vision 9865–9874.

Kingma, D. P., and Ba, J. (2014). Adam: A method for stochastic optimization. arXiv preprint 1412.6980.

Monk, C.S., Peltier, S.J., Wiggins, J.L., Weng, S.J., Carrasco, M., Risi, S., Lord, C. (2009). Abnormalities of intrinsic functional connectivity in autism spectrum disorders. Neuroimage 47, 764– 772. doi: 10.1016/j.neuroimage.2009.04.069

Niu, K., Guo, J., Pan, Y., Gao, X., Peng, X., Li, N., Li, H. (2020). Multichannel deep attention neural networks for the classification of autism spectrum disorder using neuroimaging and personal characteristic data. Complexity 2020. doi: 10.1155/2020/1357853

Paszke, A., Gross, S., Massa, F., Lerer, A., Bradbury, J., Chanan, G., et al. (2019). PyTorch: An imperative style, high-performance deep learning library, in Advances in Neural Information Processing Systems, eds. H. Wallach, H. Larochelle, A. Beygelzimer, F. Alché-Buc. d\ textquotesingle, E. Fox, R. Garnett (Red Hook, NY: Curran Associates, Inc.), 8024–8035.

Pedregosa, F., Varoquaux, G., Gramfort, A., Michel, V., Thirion, B., Grisel, O., Blondel, M., Prettenhofer, P., Weiss, R., Dubourg, V., Vanderplas, J. (2011). Scikit-learn: Machine learning in Python. J. Mach. Learn. Res. 12, 2825–2830.

Shen, H., Wang, L., Liu, Y., Hu, D. (2010). Discriminative analysis of resting-state functional connectivity patterns of schizophrenia using low dimensional embedding of fMRI. Neuroimage 49, 3110–3121. doi: 10.1016/j.neuroimage.2009.11.011

Supekar, K., Musen, M., Menon, V. (2009). Development of large-scale functional brain networks in children. PLoS Biol. 7, e1000157.

Yan, W., Wang, Y., Gu, S., Huang, L., Yan, F., Xia, L., Tao. Q. (2019). The domain shift problem of medical image segmentation and vendor-adaptation by Unet-GAN, Medical Image Computing and Computer Assisted Intervention – MICCAI 2019 (pp.623-631). doi :10.1007/978-3-030-32245-8_69

## References

Ktena, S.I., Parisot, S., Ferrante, E., Rajchl, M., Lee, M., Glocker, B., Rueckert, D. (2018) Metric learning with spectral graph convolutions on brain connectivity networks. NeuroImage 169, 431–442.

Loshchilov, I., and Hutterm F. (2016). Sgdr: Stochastic gradient descent with warm restarts. arXiv Preprint. 1608.03983.

